# The prevalence of bleeding disorders in women with proven endometriosis: results of a screening study

**DOI:** 10.1101/2023.03.03.23286728

**Authors:** Sarah Heynemann, Emma Johnson, Melissa Cameron, Ray Dauer, Elya Moore, Chris Barnes, Sonia R Grover

## Abstract

**Objective:** To establish the rate of bleeding disorders amongst women with confirmed endometriosis and clarify predictive features on history.

**Methods:** Eight-two women with confirmed diagnosis of endometriosis were recruited from a tertiary womens’ hospital. General bleeding tendency was evaluated with a modified internationally standardised questionnaire, and a bleeding score was calculated. Menstrual loss was evaluated by obtaining current and heaviest-ever menses profile using Pictorial Blood loss Assessment Chart(PBAC). Forty-one women satisfied criteria for a significant bleeding tendency and underwent coagulation tests including Prothrombin time(PT), activated partial thromboplastin time(aPTT), Platelet Function Analyser-100(PFA-100) and von Willebrand factor(vWF)/Factor VIII(FVIII) studies.

**Results:** The prevalence of abnormal screening haemostasis tests for the population was 17.72% (95% CI 10.04-27.94) for all tests, 8.54%(95% CI 3.50-16.80) for vWF tests, and 12.99%(95% CI 6.41-22.59) for PFA-100. Receiver Operator Characteristic curve analysis demonstrated a ‘very good’ performance of the bleeding score as a diagnostic test for haemostatic abnormalities for nulliparous women with endometriosis. Bleeding symptoms individually were not predictive. Logistic regression suggested the combination of mittleschmerz, cutaneous bleeding symptoms, heavy menstrual bleeding and prolonged bleeding from minor wounds as predictive for haemostatic abnormalities (p=0.0197).

**Conclusion:** The prevalence of abnormal bleeding tests in women with endometriosis on preliminary testing was higher than the population rate of bleeding abnormalities and thus people with histologically proven endometriosis warrant a higher index of suspicion with respect to testing for mild bleeding disorders.

Bleeding disorders in women with endometriosis

## Introduction

Endometriosis is a benign condition primarily affecting women of reproductive age. A definitive diagnosis of endometriosis is based on histopathological confirmation of suspected endometriotic biopsies taken during surgery and this remains the gold standard in clinical guidelines from many national and international societies (As-Saine, Black et al 2019). A systematic review of the correlation between histology and surgical visualization reported a ‘negative’ laparoscopy is likely to be accurate in excluding endometriosis, however a ‘positive’ laparoscopy, at which disease is visualized surgically, is less informative, hence histological confirmation is advocated(Wykes, Clark et al. 2004).

Although there is some preliminary work suggesting that a non-invasive diagnostic test may be on the horizon (Bendifallah, Suisse et al 2022), estimates of endometriosis disease prevalence are limited to hospital or surgical audits. Whilst a population prevalence rate of 10% has been proposed (Eskenazi and Warner 1997), and is generally accepted, inconsistency in case definition for endometriosis continues to be a significant concern (Koninckx, Ussia et al 2021) and rates vary depending on population selection with the highest rates in those with infertility(25-50%) or chronic pelvic pain (71-87%) (As-Saine, Black et al 2019). Identifying the lack of symptoms or cluster of present symptoms that predict for a diagnosis of endometriosis remains a challenge (Surrey, Carter et al 2017) as does the lack of correlation between severity of symptoms and severity of endometriosis (Agarwal, Chapron et al 2019).

Although retrograde menstruation is considered a physiological process present in close to 100% of women undergoing laparoscopy at the time of menses (Halme, Hammond et al. 1984), the challenge is understanding why endometriosis does not occur in all women. Although ‘implantation’ is the prevailing theory for the pathogenesis of peritoneal endometriosis, other factors including immune dysfunction, oxidative stress and stem cells may all contribute (Burney and Guidice 2012). Observations to support this include the higher prevalence of retrograde menstruation in baboons and women with spontaneous endometriosis than those with a normal pelvis and experimental implantation (D’Hooghe and Debrock 2002). In studies on the risk factors for endometriosis, a consistent finding is that exposure to menstruation, with earlier age of menarche, shorter cycle duration (Missmer and Cramer 2003), and longer and heavier menses(Cramer, Wilson et al. 1986; Vercellini, De Giorgi et al. 1997) are all associated with an increased risk of disease, whilst there is a decreased risk with increasing parity (Eskenazi and Warner 1997).

Given the evidence that the volume of retrograde menstrual loss (D’Hooghe and Debrock 2002) appears to relate to endometriosis risk, it may be more likely that women who are unable to clear this magnified potential stimulus for disease (Cramer and Missmer 2002) are at higher risk. Heavy and prolonged menstrual bleeding are common symptoms amongst women with bleeding disorders, and a higher prevalence of bleeding disorders amongst women with heavy menstrual bleeding (HMB) compared to the general population has been reported, with a 13% rate for vonWillebrand Disease(vWD) (Shankar, Lee et al. 2004). For women with vWD, menses may pose a considerable problem and impact on quality of life (Kirtava, Drews et al. 2003; Lee and Kadir 2005; James, Ragni et al. 2006); additionally, bleeding may occur during pregnancy, postpartum, and at ovulation (Kouides, Phatak et al. 2000; James 2005). Although the presence of endometriosis was not histologically verified, in a survey of women with vWD, 30% of women reported having been told at some stage by a doctor that they had endometriosis (Kirtava, Drews et al. 2003). Despite the epidemiological evidence suggesting an affiliation between endometriosis and bleeding disorders (Vercellini, De Giorgi et al. 1997) and a report on a clinical observation(Mitri and Casper 2015), there have been no studies to date to confirm this.

Although the reported rate of vWD in the general population has been 1% (Rodeghiero, Castaman et al. 1987; Werner, Broxson et al. 1993), both of these epidemiological studies did not focus on medically significant or symptomatic bleeding, and with subsequent follow-up many of these individuals did not develop symptomatic or significant bleeding. A primary care study exploring the prevalence of symptomatic vWD nevertheless concluded that the population rate was at least one in a thousand (Bowman, Hopman et al 2010).

Platelet function disorders are thought to be of a similar frequency to vWD (Lusher 1999). Other bleeding disorders are rare and have prevalence rates in the order of 1 in 10,000 for Factor IX (Peyvandi, Duga et al. 2002), to 1 in 1,000,000 for Factor XI (Mannucci and Tuddenham 2001; Peyvandi, Duga et al. 2002), although an increased prevalence occurs in populations of Ashkenazi Jewish descent(Peyvandi, Duga et al. 2002).

Despite bleeding disorders being recognized as a significant predisposing factor for HMB in premenopausal women, identification of these women has remained low. A survey in 2006 of obstetricians and gynaecologists revealed that only 2% of gynaecologists would consider investigating a 35 year-old woman with HMB for a mild bleeding disorder (Chi, Shiltagh et al 2006).

Identifying women with a heightened bleeding tendency is challenging. Clinical determination of ‘significant’ or ‘excessive’ bleeding tendency is fraught by non-specific symptoms and heterogeneity in clinical manifestations (Greaves and Watson 2007), with ‘haemorrhagic symptoms’, real or perceived, present in apparently normal individuals (Sadler 2003; Rodeghiero, Tosetto et al. 2007; Rodeghiero 2008). A structured bleeding history has been shown to have some utility (Sramek, Eikenboom et al. 1995; Rodeghiero, Castaman et al. 2005; Tosetto, Rodeghiero et al. 2006; Philipp, Faiz et al. 2008) although the criteria for concluding that there is a bleeding disorder influences the degree of predictiveness or utility. In the widely accepted study by Rodegheiro, specificity of a bleeding history questionnaire was >90% and sensitivity 80% (Rodeghiero, Castaman et al. 2005). An additional tool that has been used for attempting to identify women with heavy menses is the pictorial blood loss assessment chart (PBAC) (Higham, O’Brien et al. 1990), with the combination (ie., questionnaire plus PBAC) used by some – either stepwise to identify women for further screening (Kadir, Economides et al. 1998), or as part of a multi-component screening tool (Philipp, Faiz et al. 2008).

Thus this pilot study aimed to investigate the rate of bleeding disorders amongst women with proven endometriosis, and clarify predictive features on clinical history by utilizing both a standardised questionnaire and the PBAC.

## Methodology

Women aged 16-50 years with a history of histologically confirmed endometriosis were recruited from the endosurgery and gynaecology clinics of the Mercy Hospital for Women (MHW) and private rooms of the clinicians involved between October 2008 and March 2009. In the absence of histology, surgical visualization and/or photographs of distinctive endometriomas with obliteration of the Pouch of Douglas were considered sufficient evidence of endometriosis. Women with an intellectual disability or language difficulties, or a previously diagnosed bleeding disorder were excluded. Women with adenomyosis were only included if they also had proven endometriosis.

Following recruitment and written consent, general and menstrual bleeding tendency were evaluated using a standardized questionnaire and PBACs administered by SH or EJ. General bleeding tendency was investigated using a modified version of the standardized questionnaire originally proposed by Rodeghiero et al. Each symptom could be considered ‘trivial’ or ‘significant’, and details were agreed upon via research team consensus prior to data collection following specialist haematological and gynaecological input. Symptoms included epistaxis, cutaneous bleeding, oral cavity bleeding, gastrointestinal bleeding, postpartum haemorrhage, muscle haematomas, prolonged bleeding from minor wounds/after teeth extraction, and HMB; with 0-3 points allocated according to symptom severity. Modification to this questionnaire included the addition of mittleschmerz (midcycle, ovulation related pain) (Kouides, Phatak et al. 2000; James 2005; Lee, Chi et al. 2006) and family history of excessive bleeding(Sramek, Eikenboom et al. 1995; Jayasinghe, Moore et al. 2005) given research subsequent to development of the original questionnaire by Rodeghiero et al (2005) noting these to be discriminatory symptoms for bleeding disorders. A summative bleeding score was calculated based on questionnaire responses. Women were identified as warranting haemostatic testing if they had a score >4 for nulliparous women, >5 for parous women, or the presence of ≥ 3 symptoms (“presence’ was defined by achievement of ≥1 point, or ≥2 points for family history). Information regarding current medications, history of other gynaecological conditions, anaemia and liver disease was also recorded. Modification to the bleeding score was undertaken to promote high sensitivity of the tool to maximise likelihood of identifying women with clinical history suspicious for a bleeding disorder for subsequent haemostatic testing, with lowering of the score for women who had not experienced the haemostatic challenge of childbirth. The PBAC was administered to evaluate both current (if applicable) and ‘heaviest ever’ menstrual profile. A score of ≥100points for the PBAC was considered evidence of HMB as has been used in the original study by Higham et al (1990).

Women who satisfied the described criteria underwent testing for possible vWD and platelet dysfunction. Blood was taken by experienced phlebotomists and all test processing except for the PFA-100® (Platelet Function Analyser-100) performed by Austin Hospital Pathology. Where possible the day of the woman’s menstrual cycle was recorded. Haemostatic evaluation included prothrombin time (PT) (using Hemosil RecombiPlasTin ®. Instrumentation Laboratory, Lexington, Ma, USA); Activated Partial Thromboplastin Time (APTT) (performed using Triniclot HS, which uses purified phospholipids containing micronised silica as the activator, Trinity Biotech, USA); von Willebrand Factor Antigen (vWF:Ag) (performed using the HemosIL™ Von Willebrand Factor Antigen kit. Instrumentation Laboratory, Lexington, Ma, USA); vWF Ristocetin Cofactor (vWF:RCo) (performed using stabilized platelets and ristocetin in a lyophilized form, from Dade Behring/Siemens, Inc Newark, DE, USA); vWF:collagen binding assay (vWF:CBA) (performed using an ELISA based method from Life Therapeutics. Life Therapeutics, Frenchs Forest, NSW, Australia); Factor VIII:C. (Factor VIII activity was performed using a one stage APTT based method. Factor VIII deficient plasma was obtained from Dade Behring/Siemens, Inc Newark, DE, USA). Platelet function was assessed using PFA-100® (Dade Behring/Siemens, Inc Newark, DE, USA), utilising both Collagen Epinephrine (C-EPI) and Collagen Adenosine Diphosphate (C-ADP) cartridges using 3.2% citrated whole blood between 30 minutes and 4 hours after blood draw as recommended (Hayward, Harrison et al. 2006).

Results for vWF:RCo were considered abnormal if <50%; vWF:Ag and vWF:CB results between 50-55% were considered borderline and in the context of this screening study, abnormal. The haematocrit value had to be between 0.35 – 0.50, and platelets between 150-500 × 10^9^/L for specimens to be included for PFA-100 ® testing. Reference ranges for PFA-100 ® C-EPI Closure Time (CT) ≤ 170 seconds, C-ADP CT ≤115secs. Where possible, testing was rescheduled if patients reported having used a list of common nonsteroidal anti-inflammatory drugs (NSAIDs) or aspirin in the 2 weeks prior to PFA-100 ®, otherwise the number of days since last NSAID usage was recorded.

Women who returned an ‘abnormal’ result for any of the initial tests performed, according to stated reference ranges, were included for analysis in this study. For the purpose of prevalence estimates, women who did not satisfy criteria for increased bleeding tendency and were not tested were assumed to have normal haemostasis.

### Endometriosis

Clinical notes and photographs available from the operation/s at which endometriosis was diagnosed were reviewed by a single gynaecologist with an interest in endometriosis.

### Statistical analysis

Questionnaire and PBAC data were entered into an Excel database, blinded to blood test results, and analysis was performed using STATA™ 8.2. P<0.05 was considered statistically significant.

Prevalence rates of haemostatic abnormalities were calculated separately for vWF/FVIII, PFA-100® and all tests combined. Spearman Rank Correlation was performed to determine if non-parametric continuous variables such as bleeding score, number of symptoms, PBAC score and age were independent of each other. Mann-Whitney tests were performed to examine the effect of parity status on distribution of bleeding score, number of symptoms and PBAC scores; and to determine if PBAC scores differed between women with ‘abnormal’ and ‘normal’ haemostasis results. Wilcoxon signed rank test was performed to determine if ‘current’ and ‘heaviest ever’ PBAC scores differed significantly. Receiver Operator Characteristic (ROC) curve analysis was performed to determine the individual value of symptoms, bleeding score, number of symptoms, and PBAC as diagnostic tests for initial haemostatic abnormalities. ROC curves display the compromise between sensitivity and specificity at different cut-offs of a diagnostic test (McPherson and Pincus 2007). Area under the curve (AUC) can be calculated as a summary measure, with <0.7 considered ‘poor’ and >0.9 considered ‘excellent’ (McPherson and Pincus 2007). Step-wise backward selection grouped linear logistic regression was performed to determine the most predictive combination of symptoms.

### Ethical approval

Ethical approval was granted by MHW Human Research and Ethics Committee for the project (R08/15).

## Results

Between October 2008 and March 2009, 105 women with endometriosis were recruited. Of these 23 were subsequently excluded from the study with 9 failing to undergo haemostatic testing despite having scores suggestive of a significant bleeding tendency, and a further 14 due to an inability to confirm their diagnosis of endometriosis. Of the resultant 82 participants, the vast majority reported that their endometriosis had been diagnosed following investigation for pain (dysmenorrhoea/generalised pelvic pain /dyspareunia /mittleschmerz). Three women reported infertility as a cause for investigation and 1 woman infertility and dysmenorrhoea. General population descriptors are shown in Table 1.

**Table 1.**
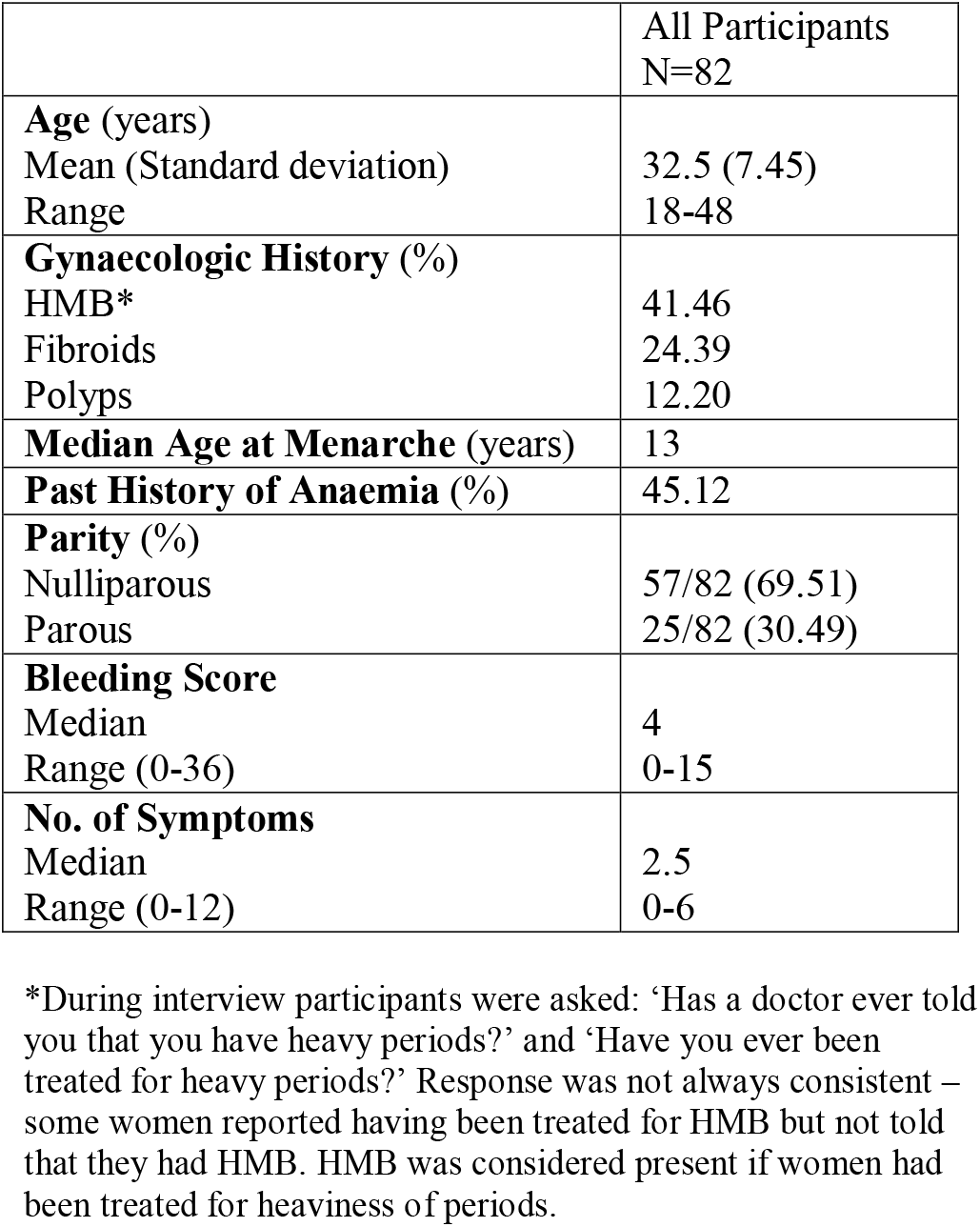
Population characteristics

Forty-one women underwent haemostatic testing following satisfaction of described criteria.

One woman had a prolonged APTT, and all PT tests were normal. At least one vWF/FVIII test was found to be abnormal in 7/82 women giving a prevalence of 8.54% for abnormal haemostatic tests (95%CI 3.50%-16.80%). Excluding 3 women with prolonged PFA-100 ® results in the context of having taken NSAIDs ≤ 14 days prior to testing, and 2 women who did not have PFA-100® tests performed, 10/77 women had at least one abnormal PFA-100® result giving a prevalence of 12.99% for abnormal PFA-100 ® results (95%CI 6.41-22.59%). Prolonged PFA-100® results were observed in several women in the context of NSAID use ≤ 14 days prior to testing, low haemotocrit and/or low platelet count, although in other women with the same factors, the results were normal.

Therefore 14/79 women had at least one abnormal test, giving a prevalence of 17.72% abnormal haemostatic tests (95% CI 10.04-27.94%). This, and all subsequent references to the overall prevalence for all tests combined excludes: 2 women for which PFA-100® was not performed and 1 woman whose only abnormal result was prolonged PFA-100® in the setting of having used NSAIDs ≤14 days prior to testing.

### Bleeding score, number of symptoms, individual bleeding symptoms

Spearman Rank Correlation provided strong evidence for a strongly positive relationship between bleeding score and number of symptoms, and a weakly positive relationship between bleeding score and PBAC scores. Age appeared to be independent of each of bleeding score, number of symptoms, and PBAC scores (Table 2).

**Table 2.** Spearman Rank Correlation analysis

|  | P value | Spearman Rank Correlation coefficient |
| --- | --- | --- |
| **Age and:** |  |  |
| bleeding score | 0.701 |  |
| number of symptoms | 0.914 |  |
| PBAC 'current' score | 0.682 |  |
| PBAC 'heaviest ever' score | 0.858 |  |
| **Bleeding score and:** |  |  |
| number of symptoms | **<0.0001** | 0.939 |
| PBAC current score | **0.002** | 0.374 |
| PBAC 'heaviest ever' score | **0.001** | 0.367 |
| **PBAC 'current' score and:** |  |  |
| bleeding score | **0.0021** | 0.3744 |
| number of symptoms | **0.0039** | 0.3537 |
| **PBAC 'heaviest ever' score and:** |  |  |
| bleeding score | **0.0007** | 0.3673 |
| number of symptoms | **0.0003** | 0.3946 |

There did not appear to be evidence of a significant difference in age distribution between the women with normal haemostasis and abnormal test/s(p=0.792), including when Mann-Whitney tests were performed separately for vWF/FVIII (p=0.601) and PFA-100® (p=0.710). Parity had no effect on each of bleeding score, number of symptoms, and PBAC scores when examined via the Mann-Whitney test. Likewise PBAC score distribution of ‘current’ score versus heaviest ever’ score had no impact on normal versus abnormal tests for vWF tests, or all tests combined.

With respect to the clinical indicators of general bleeding tendency, the parous and nulliparous women were analysed separately, since different cut-off were used. Utilising the ROC curves to evaluate the value of the bleeding score in nulliparous women (70% of the population), the bleeding score demonstrated promise as a tool to discriminate between those with normal and abnormal haemostasis, with very good Area Under Curve (AUC) point estimates (≥0.80) for all tests, vWF/FVIII and PFA-100® abnormalities. Bleeding score had a poor performance amongst parous women. Number of symptoms had a fair performance for all tests (AUC = 0.779 (95% CI 0.68-0.87)), vWF/FVIII tests (AUC 0.76 (95% CI 0.65-0.88)) and PFA-100® (AUC 0.77(95%CI0.67-0.87).

For vWF/FVIII abnormalities individual symptoms performed poorly, including several AUC point estimates around 0.5, which is considered a diagnostic test failure. For PFA-100®, HMB and cutaneous symptoms had good AUC point estimates, however 95% CI were quite wide, and the lower limit surrounded 0.5 for 58 women with cutaneous symptoms. For all tests, HMB had a good point estimate for AUC, however 95% CI were quite wide. In the 25 parous participants PPH generated a good AUC point estimate for PFA-100® and all tests, however 95% CI were very wide and poor.

Using a ≥100 point threshold for ‘current’ and ‘heaviest ever’ PBAC scores respectively, 53.85% and 76.54% of women had a score suggestive of HMB (Higham, O’Brien et al. 1990). Spearman Rank Correlation provided strong evidence of a weak-moderate positive relationship between PBAC ‘current’, ‘heaviest ever’ and bleeding score and number of symptoms. There was a significant difference in median PBAC score between ‘current’ and ‘heaviest ever’ PBAC, regardless of whether women were taking medications which may have affected menstrual flow (combined oral contraceptive pill, levonorgestrel intrauterine system, gonadotrophin releasing hormone analogues, tranexamic acid) at time of ‘current’ PBAC or not. PBAC score distribution did not appear to differ significantly between women with normal and abnormal haemostatic result/s, except current PBAC for PFA-100® results (p=0.0415). As a diagnostic test to discriminate between women with normal and abnormal haemostatic result/s both ‘current’ and ‘heaviest ever’ PBAC had a poor performance, and only ‘current’ PBAC had a good point estimate for ROC AUC, although 95% CI were wide and poor (0.71, 0.56-0.87).

## Discussion

Studies have shown that women with HMB have a higher rate of mild bleeding disorders than the background population rate (Shankar, Lee et al. 2004). Although there is work that demonstrates that HMB is a risk factor for endometriosis (Vercellini, De Giorgi et al. 1997), this study provides the first indication of the prevalence of bleeding test abnormalities in women who have a confirmed diagnosis of endometriosis. This finding is consistent with the epidemiological studies demonstrating an association between endometriosis and factors predisposing to greater exposure to menses (for example HMB) and the potential opportunity for increased retrograde menstruation (Cramer, Wilson et al 1986; Vercellini, De Giorgi et al. 1997; Missmer 2003), with subsequent implantation as endometriotic deposits (D’Hooghe and Debrock 2002) in accordance with Sampson’s hypothesis.

The overall prevalence of 17.72% of initial laboratory haemostatic abnormalities in the sample, is lower than may be expected. Previously, the only other suggestion of an increased rate of bleeding disorders amongst this population, has been the unverified self reported 30% rate of endometriosis reported in a survey of women with vWD (Kirtava, Drews et al. 2003). In contrast, particular effort was undertaken in this study to have either histological proof or very clear supporting visual evidence of endometriosis (not simply “pigmented spots”).

In earlier studies, the prevalence of definitive bleeding disorders (compared to initial abnormal haemostatic screening tests) was reduced following specialist evaluation, as occurred in Kadir’s study (Kadir, Economides et al. 1998). In Kadir’s study, only 65% of those who initially had results suggestive of vWD were defined as having the disorder on follow-up and repeat testing. Regardless, the rate of mild bleeding disorders appears to be greater amongst women with endometriosis than the rate in the general population.

The detection of an underlying bleeding abnormality in a woman with endometriosis is pertinent to clinical management on several levels. The use of NSAIDs to manage dysmenorrhoea/pelvic pain amongst women with endometriosis is common. Although in healthy populations NSAIDs have been shown to ameliorate HMB compared to placebo (Lethaby, Augood et al 2007), in women with bleeding disorders they may be counter-productive in addressing HMB (Lee, Chi et al 2006; Siegel and Kouides 2002).

Further prospective study is required to determine if treatment strategies aimed at reducing menstrual blood loss would be beneficial in this group. Epidemiological data suggests that reduced menstrual loss associated with increased parity (Eskenazi and Warner 1997) and the use of the oral contraceptive pill appear to reduce the risk for endometriosis (Vercellini, Eskenazi et al 2011).There is evidence from small studies to suggest this may be effective in adolescents where the rate of endometriosis in adolescents with dysmenorrhea was vastly lower than expected in a clinical setting where menstrual loss had been reduced with the use of tranexamic acid or hormonal menstrual management prior to considering a laparoscopy to investigate their pain (Sachedina, Abu Bakar et al 2020). Long term follow-up of this cohort also revealed a lower than expected rate of endometriosis (Knox, Ong et al 2019).

There are some significant methodological limitations to this pilot study which warrant noting regarding the detection of bleeding disorders.

Firstly, with respect to participants recruited, limitations relate to loss to follow-up of 9 women who satisfied the criteria for ‘significant’ bleeding tendency however did not undergo haemostatic testing and were subsequently excluded, which may have falsely limited the prevalence of haemostatic abnormalities. A further 14 were excluded due to an inability to confirm their endometriosis diagnosis (with their operative procedure performed at other hospitals and despite efforts, their notes could not be traced).

Secondly, there were limitations with respect to the strength of the haemostasis testing approach. Funding limitations restricted our capacity to undertake repeat haematological testing amongst both the women who underwent initial haemostatic testing and returned abnormal results, as well as those with initial apparently normal results. However, further and repeat testing were beyond the scope of this pilot study. It is it is acknowledged that a formal diagnosis (and implications of labelling) of a bleeding disorder for a patient requires repeat testing and clinical evaluation by specialist haematological services. Women in the study who returned abnormal initial haemostatic results, according to the criteria described, were referred on for further haematological evaluation. For women in this study who did not satisfy the described criteria for suspicion of increased bleeding testing (following data acquired via the questionnaire and PBAC responses) haemostatic testing was not performed. This was both pragmatic due to funding limitations as well as recognizing that, for some, in the absence of significant bleeding history there may have been lack of enthusiasm to undergo testing and this may have adversely affected recruitment to this pilot study. Whilst measures to optimize the sensitivity of the screening tools for suspicious bleeding history were applied in this study as described, it is possible that women who were not tested may nonetheless have also returned abnormal haemostatic tests had they been tested (hence the prevalence estimates for abnormal haemostatic testing may be conservative). Future research involving testing of all women with endometriosis, whether they have a suspicious bleeding history or not, as well as repeat and longitudinal testing, and with specialist haematological input will be critical to confirm the results of this hypothesis-generating pilot study.

Recognition of the abovementioned methodological limitations lead to the failure to submit this paper for publication at time of completion. More recently, realization by the senior author who has an ongoing interest in this area that there have still been no studies to clarify the relationship between endometriosis and mild bleeding disorders, despite the significant passage of time since this study was undertaken, has lead to the decision to submit the paper now. This is in recognition that despite the data having a number of limitations as described with candour, it represents the only effort to tackle the question regarding the potential relationship between mild bleeding disorders and endometriosis. Acknowledgement of this as a potential contributor to endometriosis may offer some women further options for treatment, the avoidance of potential surgical risks as well as the possibility of earlier intervention. As such, we would applaud future research with a larger participant sample, and which overcome the haematological testing limitations of this pilot study described, to further investigate the hypothesis-generating data reported here which may suggest an increased bleeding tendency exists amongst some women with endometriosis.

## Data Availability

All data produced in the present study are available upon reasonable request to the authors

